# A Mathematical Model and Optimal Control for Listeriosis Disease from Ready-to-Eat Food Products

**DOI:** 10.1101/2020.10.11.20210856

**Authors:** Williams Chukwu, Farai Nyabadza

## Abstract

Ready-to-eat food (RTE) are foods that are intended by the producers for direct human consumption without the need for further preparation. The primary source of human Listeriosis is mainly through ingestion of contaminated RTE food products. Thus, implementing control strategies for Listeriosis infectious disease is vital for its management and eradication. In the present study, a deterministic model of Listeriosis disease transmission dynamics with control measures was analyzed. We assumed that humans are infected with Listeriosis either through ingestion of contaminated food products or directly with *Listeria Monocytogenes* in their environment. Equilibrium points of the model in the absence of control measures were determined, and their local asymptotic stability established. We formulate an optimal control problem and analytically give sufficient conditions for the optimality and the transversality conditions for the model with controls. Numerical simulations of the optimal control strategies were performed to illustrate the results. The numerical findings suggest that constant implementation of the joint optimal control measures throughout the modelling time will be more efficacious in controlling or reducing the Listeriosis disease. The results of this study can be used as baseline measures in controlling Listeriosis disease from ready-to-eat food products.

## 1 Introduction

Human Listeriosis is a zoonotic disease with a low incidence rate, but with a high mortality rate of those sickened with the infection globally Bennion et al. (2008). The disease poses a huge public health concern, and as a result there is need to develop strategies to combat any outbreak of the disease. Humans are infected with Listeriosis through consumption of Listeria contaminated food products or directly from the environment by acquiring the pathogen, *Listeria monocytogenes* (*L. monocytogenes*), due to improper hygiene Hu et al. (2016); WHO (2020). In order to diagnose human Listeriosis, the active bodies responsible for food-borne diseases surveillance such as the National Institute of Communicable Disease (NICD) South Africa, Center for Disease Control (CDC) USA, and World Health Organization (WHO) obtain their results through laboratory-confirmed cases. Listeriosis is usually diagnosed when a bacterial culture grows *L. monocytogenes* from body tissue or fluid, such as blood, spinal fluid, or the placenta CDC (2020). The incubation period is from 1 to 21 days, but in some patients it takes up to 90 days after the first exposure to the bacteria before it is diagnosed CDC (2020). Symptoms of infection include: fever, flu-like symptoms, nausea, diarrhoea, fatigue, headache, stiff neck, convulsion, loss of balance, and muscle aches. Furthermore, in pregnant woman, it can cause miscarriages, stillbirths, premature delivery, or life-threatening infections to the unborn babies. Human Listeria infections can be treated with *β*-lactam antibiotic, normally ampicillin Almudena and Payeras-Cifre (2014). However, upon any outbreak, some preventive measures such as the recall of contaminated food products, factory workers practicing proper hygiene, educational campaign programs can be implemented to control the disease.

Mathematical models improve our understanding of pathogen dynamics by providing a theoretical framework in which factors affecting transmission and control of the pathogens can be explicitly considered Lanzas et al. (2011). The purpose of modelling disease epidemics is to provide a rational basis for policies designed to control the spread of disease outbreaks. This includes practical optimal strategies in models that allow the assessment of the interventions instituted by public health authorities. Optimal control is a powerful mathematical tool in decision making which involves employing appropriate strategies to eradicate epidemics from the population Makinde and Okosun (2011). These decisions include; determining the proportion of the population that should be treated as time evolves in any given epidemic in order to minimize both the number of infections in the population and the cost of the treatment strategy implementation Omondi et al. (2018).

Recently, mathematical models have been used to study the transmission dynamics of Listeriosis (see for example Stout et al. (2020); Otoo et al. (2020); Osman et al. (2018); Witbooi etal. (2020); Chukwu and Nyabadza (2020)). In particular, Otoo et al. (2020) studied the optimal control of Listriosis with system of equations describing the disease transmission in both human and animal populations. Further, none of these models considers the case of optimal control strategies to control Listeriosis in the human population by ingestion of contaminated ready-to-eat food products. The objective of this study is to develop and analyse an optimal control model of Listeriosis foodborne disease (FBD) from contaminated ready-to-eat food products. The optimal control strategies to be implemented are in the form of recalls of contaminated foods products, boosting the immune system and educational campaigns.

The rest of the manuscript is organized as follows; in the next section, we give the formulation of the mathematical model, as well as the underlying assumptions. Section 3 presents the model properties and analysis without controls. In Section 4, the definition of permissible optimal controls is given with corresponding optimality system. Numerical simulation results are presented in Section 5 and Section 6 concludes the study.

## 2 Mathematical Model

The mathematical model comprises of; the human population, manufactured food products, and the bacteria (*L. monocytogenes*) population in the environment. The total human population is divided into three standard epidemiological classes of susceptible *S*(*t*), infected with *L. monocytogenes I*(*t*), and the recovered *R*(*t*), at any time *t*. The human population thus follows the standard *SIR* model with the total human population *N* (*t*) given by

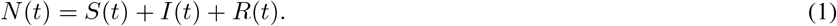

The susceptible humans grow at a rate proportional to the size of the human population, *μ*_*h*_*N* (*t*) with *μ*_*h*_ also the mortality rate of each of the human population. Susceptible humans are infected by consuming contaminated food and by Listeria from the environment at a rate Λ_*h*_ defined by Λ_*h*_ = *ω*_1_*F*_*c*_ + *ω*_2_*L*_*m*_ where *ω*_1_ and *ω*_2_ are the effective contact rates (i.e the contacts that will result in infections) for susceptible humans with contaminated food and *L. monocytogenes* respectively. Upon infection, the susceptible humans become infectious and join the compartment *I*. Once infected humans recover at a rate *α* and recovery is assumed to be with temporary immunity. Thus, recovered humans become susceptible again at rate *ρ*_*h*_ Swaminathan and Gerner-Smidt (2007). We let *L*_*m*_ represent *L. monocytogenes* with a net growth rate *r*_*l*_, and a carrying capacity 0 ≤ *κ*_*m*_ ≤ 1. The bacteria is assumed to grow logistically so that 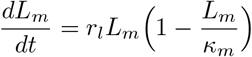. Food products are divided into two: we have uncontaminated food products *F*_*u*_ and contaminated food products *F*_*c*_, with the total food products *F* = *F*_*u*_ + *F*_*c*_. We assume a constant production rate *μ*_*f*_ of uncontaminated food with the production assumed to produce uncontaminated food. Uncontaminated food is thus contaminated at a rate Λ_*f*_ through bacteria from the environment and contaminated food in the factory’s handling as distribution processes. Here, Λ_*f*_ = *ω*_2_*L*_*m*_ + *ω*_3_*F*_*c*_ with *ω*_2_ and *ω*_3_ been the effective contact rates of the bacteria and contaminations of uncontaminated food caused contaminated food products respectively. All food products are subject to a removal rate *μ*_*f*_. We combine the model description, assumptions, model parameters, and Figure 1, which yields the following system of equations:

**Figure 1.**
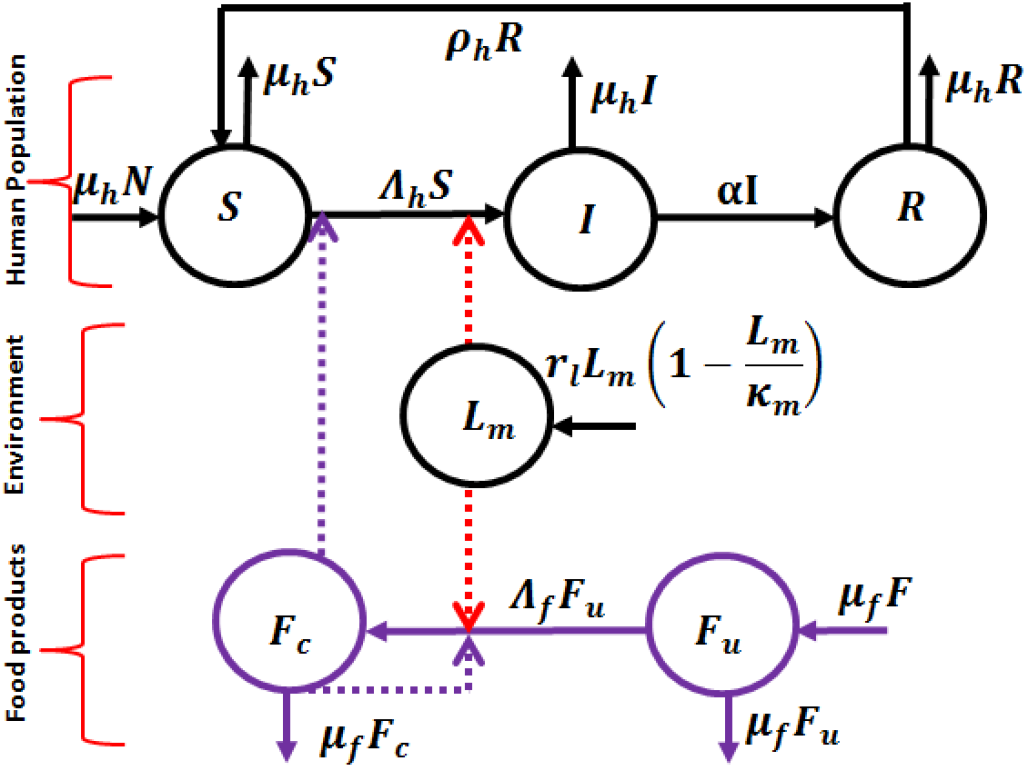
The flow chart describing Listeriosis disease transmissions dynamics within the human population from food products and *L. monocytogenes* in the environment. The solid lines indicates transitions from one compartment to another, while the dotted lines represent the influence of the other compartments on the transitions.

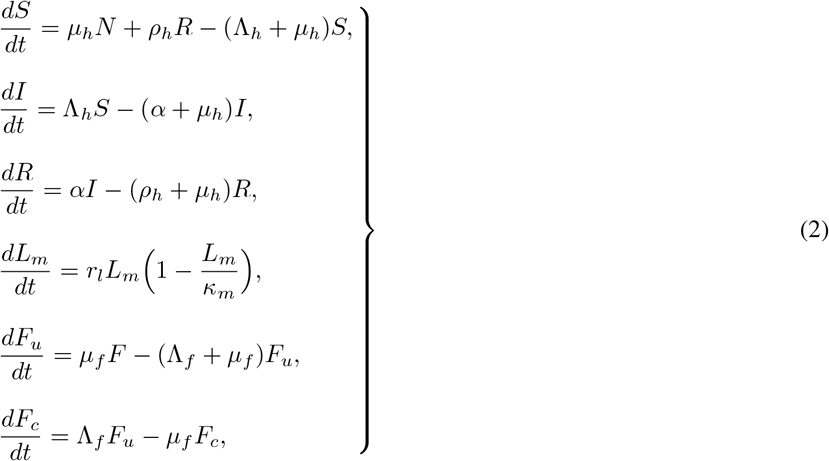

which describes the Listeriosis epidemic. We substitute *R*(*t*) = *N* (*t*) − *S*(*t*) − *I*(*t*) from (1), and transform (2) into a dimensionless systems by setting

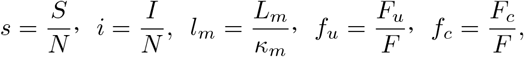

to obtain;

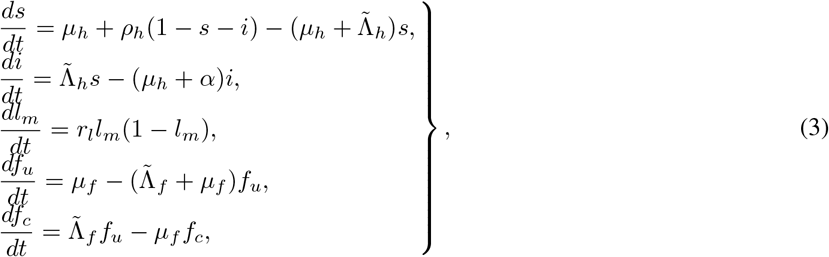

where 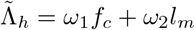, and 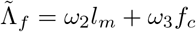, the contamination rates 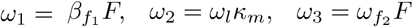, and the system (3) is subject to non-negative initial conditions

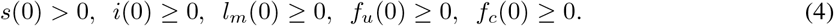

All model parameters for equations (3), are positive within the modelling time besides the net growth rate denoted by *r*_*l*_.

## 3 Dynamical Properties and Model Analysis

### 3.1 Feasible region and non-negativity of solutions

In this section we show that the solutions to equations (3) exists, are non-negative, and are boundeded in the region Ω for all *t* > 0. Assume that Ω is the biological meaningful region for model equations (3) contained in 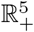. The positivity of the solutions is governed by the following theorem.

#### Theorem 1

*Model system (3) are contained in the region* Ω 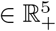, *which is given by* 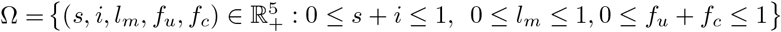, *for the initial conditions (4) in* Ω.

: The total change in human population from the first and second equations of (3) is given by the differential inequality;

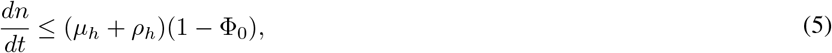

for *n* = *s* + *i* ≤ 1, and setting Φ_0_ = *s* + *i*. The solution of (5) gives

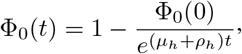

with Φ_0_(*t*) having an upper bound of 1 as 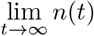 given that Φ_0_(0) ≤ 1. We consider the bacteria population in the environment given by:

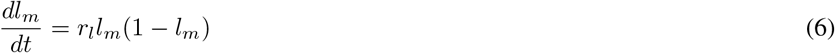

from the third equation of (3). Solving the first order equation (6) gives

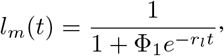

where Φ_1_ is a constant. Hence 0 ≤ *l*_*m*_ ≤ 1. The growth of *L. monocytogene* is thus bounded. Research shows that it adapts in adverse conditions events at refrigeration temperatures. This makes it easier for it to cause contamination of food products either in the factory or vegetation, there by causing Listeriosis in the human population via ingestion of contaminated foods, mostly ready-to-eat foods. Also, the total change in amount of manufactured food products from the fourth and fifth equation of (3) is given by

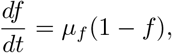

whose solution is given by

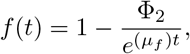

where Φ_2_ is a constant. We note that 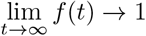. Hence, solutions of model equations (3) exists, and are biologically meaningful, bounded, and remains in Ω for *t* > ∞. We apply Theorem 2 to show, that model system (3) has non-negative solutions. We state the theorem as follows

#### Theorem 2

*For each non-negative initial conditions (4), the solutions of model equations (3)*, (*s*(*t*), *i*(*t*), *l*_*m*_(*t*), *f*_*u*_(*t*), *f*_*c*_(*t*)) *are all non-negative for t* ≥ 0.

From the first equation of the system (3), we have the differential inequality

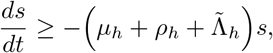

whose solutions yields

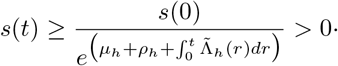

We note that 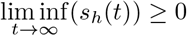, where *s*(0) > 0 is the given initial condition for the susceptible population. Hence, the solution of *s*(*t*) remains non-negative for all *t* ≥ 0. Similarly, the second equation of system (3) gives

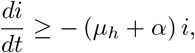

whose solutions yields 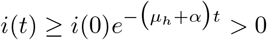, with *i*(0) as its initial condition. Similarly, it can be shown that the remaining equations of model system (3) are non-negative, that is; *l*_*m*_(*t*) > 0, *f*_*u*_(*t*) > 0, and *f*_*c*_(*t*) > 0 as *t* tends to infinity for all time *t* ≥ 0. Hence, the solutions of (3) are non-negative for all *t* ≥ 0. □

### 3.2 Steady States and their Stability Analysis

To solve for the steady states of model equation (3), we equate the right hand side to zeros as follows and obtain;

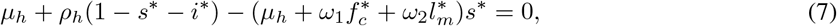

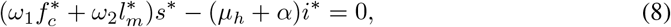

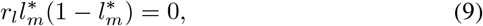

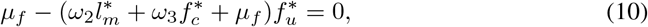

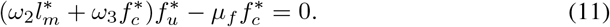

From equation (9) we have that 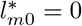 or 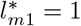. Note that, 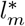 represents *l*_*m*_. If 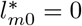, then we solve for 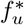 from equation (10) and obtain

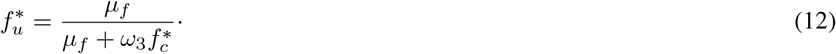

We substitute equation (12) into equation (11) and obtain

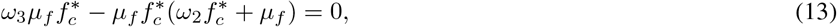

after some algebraic manipulations. Further simplifications of the expression (13) yields

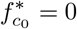

and a nonzero

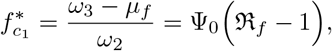

where 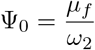 and 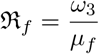.

#### Remark 1

*However, we note that* 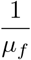 *is the duration contaminated food stays contaminated*. ℜ_*f*_ *denotes the food contamination threshold. The dimensionless quantity* ℜ_*f*_ *is the contamination of food products caused by the bacteria (Listeria), and the contaminated food* (*f*_*c*_) *during the process of food manufacturing or food preparation*. ℜ_*f*_ *connotes the basic reproduction number* ℜ_0_ *in infectious disease modeling as defined in Van den Driessche and Watmough (2002). Without loss of generality, this represents the average amount of food products that can be contaminated, and thus responsible for causing human Listeria infections*.

We have the following result on the existence of the steady state 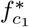.

#### Lemma 3

*The steady state* 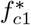 *exists subject to the existence of* ℜ_*f*_ > 1.

Thus, if 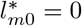, then 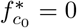 which results to 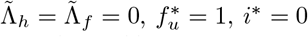, and 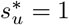. Hence we have the disease-free steady state (DFS) denoted by

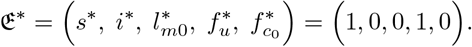

#### 3.2.1 Local stability of 𝔈*

##### Theorem 4

*The DFS of model system (3) is locally asymptotically stable whenever r*_*l*_ < 0 *and* ℜ_*f*_ < 1, *and otherwise unstable*.

: Evaluating the linearized matrix of model system (3) at 𝔈*, we have

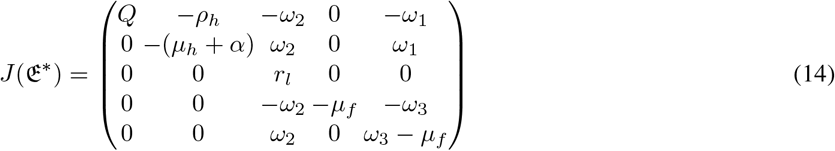

where *Q* = − (*ρ*_*h*_ + *μ*_*h*_). Hence, the eigenvalues from (14) are given as: *λ*_1_ = − (*ρ*_*h*_ + *μ*_*h*_), *λ*_2_ = − (*μ*_*h*_ + *α*), *λ*_3_ = − *μ*_*f*_, *λ*_4_ = *r*_*l*_, and *λ*_5_ = *μ*_*f*_ (ℜ_*f*_ − 1). Note that, if ℜ_*f*_ < 1, then *λ*_5_ < 0. However, if *r*_*l*_ < 0, then all the eigenvalues at 𝔈* are negatives. Then 𝔈* is locally asymptotically stable. Furthermore, if *r*_*l*_ > 0, then 𝔈* becomes unstable steady state.

The biological significance of *r*_*l*_ < 0 is that, there is decrease in the Listeria population as the total Listeria population decreases towards its maximum limit, and thus we have a logistic growth.

Also, if 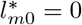, and 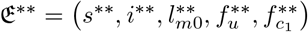, then substituting into (7)-(8) we obtain

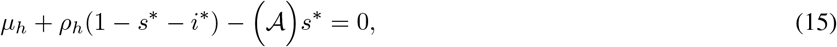

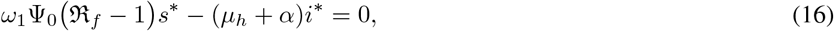

where 𝒜_0_ = *μ*_*h*_ + *ω*_1_Ψ_0_ (ℜ_*f*_ − 1). Solving (15) and (16) simultaneously for *s*\* and *i*\* gives

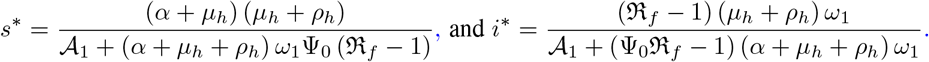

for 𝒜_1_ = (*α* + *μ*_*h*_) (*μ*_*h*_ + *ρ*_*h*_). Thus, we have the Listeriosis-free steady state (LFS) denoted by 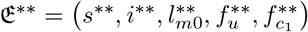 where

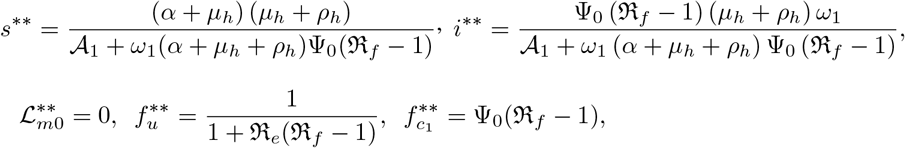

with 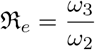. We note that 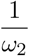 is the contamination contributed by the bacteria from the environment. The existence of 𝔈^**^ is subject to ℜ_*f*_ > 1.

#### 3.2.2 Local stability of 𝔈^**^

##### Theorem 5

*The LFS of model system (3) is locally asymptotically stable whenever r*_*l*_ < 0 *and* ℜ_*f*_ > 1, *and otherwise unstable*.

: The linearized matrix of model equations (3) evaluated at 𝔈^**^ gives

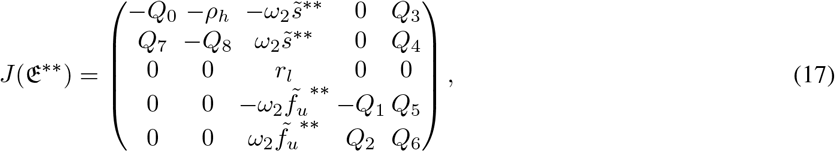

where

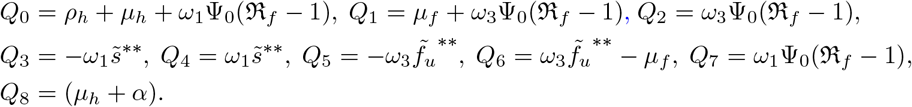

From matrix (17) the eigenvalues are: *r*_*l*_, and those obtained from the determinants of the following matrices *J*_1_(𝔈^**^) and *J*_2_(𝔈^**^) given by

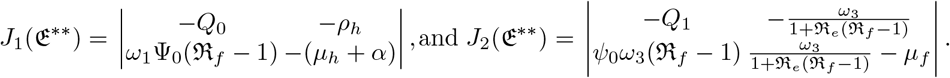

Firstly, we find the eigenvalues of *J*_1_(𝔈^**^) which are derived from the solutions of the characteristics equation

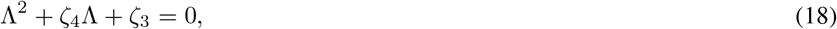

in which

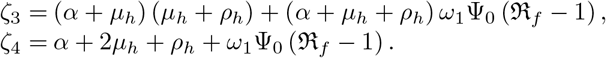

The rest eigenvalues were found from *J*_2_(𝔈^**^) given by the characteristic equation

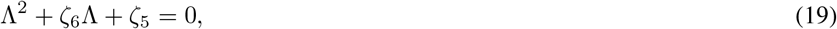

in which

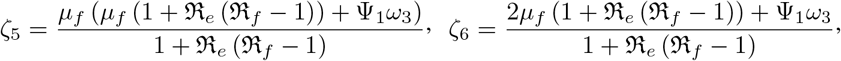

where

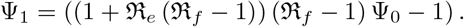

We have that *ζ*_3_, *ζ*_4_, *ζ*_5_, and *ζ*_6_ are positive if ℜ_*f*_ > 1. Hence, applying the Routh Hurwitz stability conditions, we have that all their eigenvalues of quadratic equations (18), and (19) have negative real parts. Thus, 𝔈^**^ is locally asymptotically stable if and only if *r*_*l*_ < 0, as all the eigenvalues will be negative. Note that, if *r*_*l*_ positive, we will have 𝔈^**^ becoming an unstable steady state otherwise. □

On the other hand, if *l*_*m*1_ = 1, in a similar approach we solve for *f*_*u*_ from equation (10) and obtain

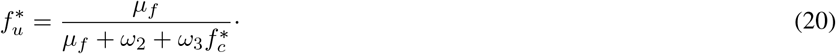

Substituting (20) into equation (11) we obtain the quadratic equation

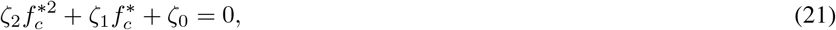

given that 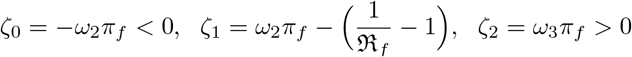. We note that *ζ*_1_ > 0 if ℜ_*f*_ > 1 and *ζ*_1_ < 0 if ℜ_*f*_ < 1. thus irrespective of the sign of *ζ*_1_, the solutions of the quadratic equation (21) 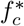 will always have one positive root say 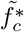. The actual value of 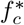 exists but cannot be determined due its intractability. Hence (7) and (8) becomes

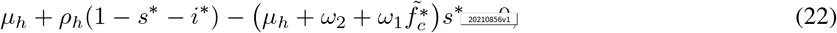

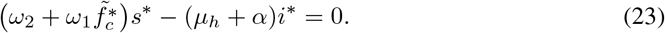

Solving (22) and (23) simultaneously for *s*\* and *i*\* gives

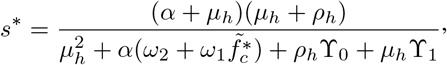

and

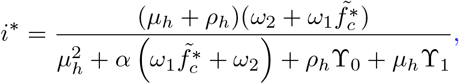

where 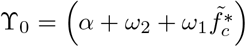 and 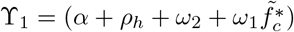. In summary, we have the Listeria endemic steady state (LES) denoted by 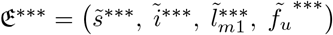 given that

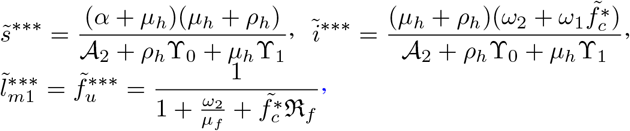

Where 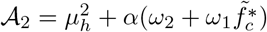.

##### Lemma 6

*The steady state* 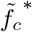 *exists subject to the existence of* ℜ_*f*_ > 1.

#### 3.2.3 Local stability of 𝔈^***^

##### Theorem 7

*The LES of model system (3) is locally asymptotically stable whenever* ℜ_*f*_ > 1, *and otherwise unstable*.

: Evaluating the linearized matrix of model equations (3) at 𝔈^***^, we have

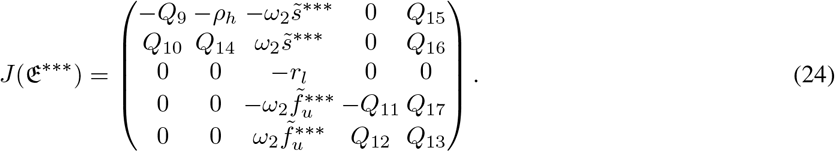

where

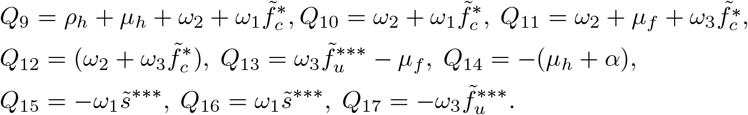

The eigenvalues of equation (24) are *λ*_6_ = −*r*_*l*_, and the solutions to the characteristics polynomial obtained

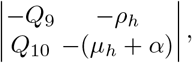

given by

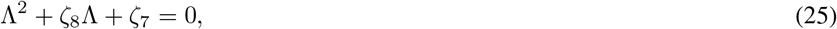

for

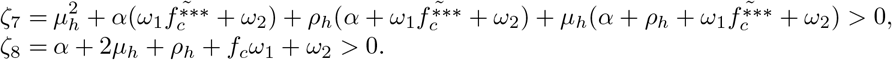

The remaining eigenvalues are determined from the determinant of

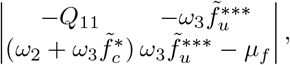

given by the characteristic equation

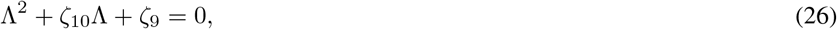

with

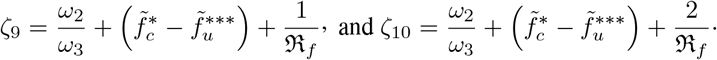

Note that *ζ*_9_ > 0, and *ζ*_10_ > 0 if and only if 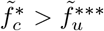. Epidemiologically, this implies that; there must be more contaminated food products than uncontaminated food products for the Listeriosis outbreak to reach an endemic state; resulting in more humans being infected with the disease. Hence, by the principle of Routh-Hurwitz stability condition, all the eigenvalues of the quadratic equations (25) and (26) have negative parts. □

### 3.3 Parameter Estimation and Sensitivity Analysis

This section is devoted to performing the sensitivity analysis of the model equations without controls. But before the sensitivity analysis, we explain how the parameter values were obtained. There are very limited resources of mathematical models and in literature based on Listeriosis disease epidemics, as a result most of the parameters values used in this work given in Table 1 were estimated except for the food product removal rate *μ*_*f*_ = 0.0099 which is from Chukwu and Nyabadza (2020). We used the fourth order Runge-Kutta numerical scheme in Matlab with a unit time step one (1) to carry out the simulations with the following initial conditions; *s*(0) = 0.6, *i*(0) = 0.3, *l*_*m*_(0) = 0.1, *f*_*u*_ = 0.7, *f*_*c*_ = 0.3, which initial conditions are chosen hypothetically for illustrative purposes and do not reflect real-life applications. With no control, the food contamination number ℜ_*f*_ is 1.5, thus, indicating the disease-free equilibrium is unstable.

**Table 1.** Parameter values used for the simulations of model without control equation (3):

| Symbols | Description | Baseline value/day |
| --- | --- | --- |
| $\mu_h$ | Natural death rate | 0.1 |
| $\alpha$ | Recovery rate | 0.0094 |
| $\rho_h$ | Lost of immunity rate | 0.09 |
| $r_l$ | Net growth rate of *L. Monocytogenes* | 0.02 |
| $\omega_1$ | Rate of human Listeriosis infection by contaminated food | 0.038 |
| $\omega_2$ | Rate of food contamination by *L. Monocytogene* | 0.002 |
| $\omega_3$ | Rate of contamination of uncontaminated food products | 0.0005 |
| $\mu_f$ | Removal rate of food products | 0.0076 |

Next, we simulate model (7)-(11) using Latin Hypercube Sampling method in Blower and Dowlatabadi (1994). This method allows us to determine which parameter of the model are more or less sensitive, and hence determine the variability for the Listeriosis disease epidemic to persist in the human population. Given the input parameters of model equations (7)-(11), the uncertainty analysis gives the model parameter output with positive, and negative partial correlation coefficients (PRCC’s) to the model system. We note that the parameters with PRCC’s > 0 will have a positive impact and those with PRCC’S < 0 will negatively impact on chosen model output. We have used a 1000 runs with a time step 1 to carry out the sensitivity analysis. The Tonardo plot Figure 2 depicts all the model parameters with their respective partial correlation coefficients. Figures 3(a) and 3(b), shows the parameters *ρ*_*h*_ and *α* are strongly positively correlated and have positive PRCC’s to the infected humans respectively, while the parameters are negatively correlated to infected humans as depicted in Figures 3(c) and 3(d). The figures show the parameters *ω*_1_, and *ω*_2_ are negatively correlated with respect to the fraction of infected humans. This results implies that; if we increase the rate of lose of immunity, fewer humans recover from Listeria infections. On the other hand, increasing *ω*_1_ and *ω*_2_ will lead to more Listeria infection in the human populations. Further, in Figure 2 we see that the removal rate of food products *μ*_*f*_ has a positive PRCC’s,while the rate of contamination of uncontaminated food, *ω*_3_, is negatively correlated. This confirms the results depicted by the food contamination threshold ℜ_*f*_ obtained in Section 3.2. Hence, increase in *μ*_*f*_ decreases ℜ_*f*_ and increase in *ω*_3_ increases the ℜ_*f*_ resulting in more human Listeriosis disease.

**Figure 2.**
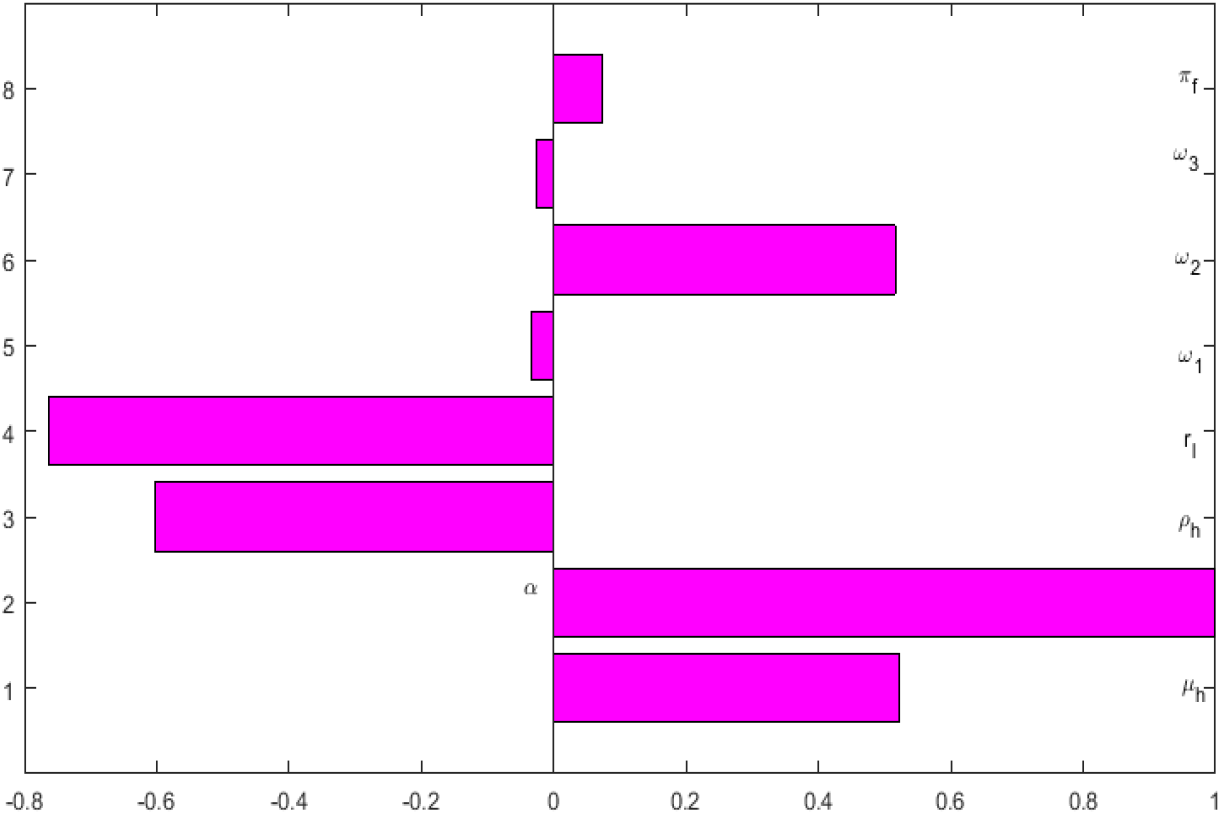
Tornado plot showing PRCC’S of all the parameters of model system (7)-(11) which are responsible for Listeriosis disease epidemics.

**Figure 3.**
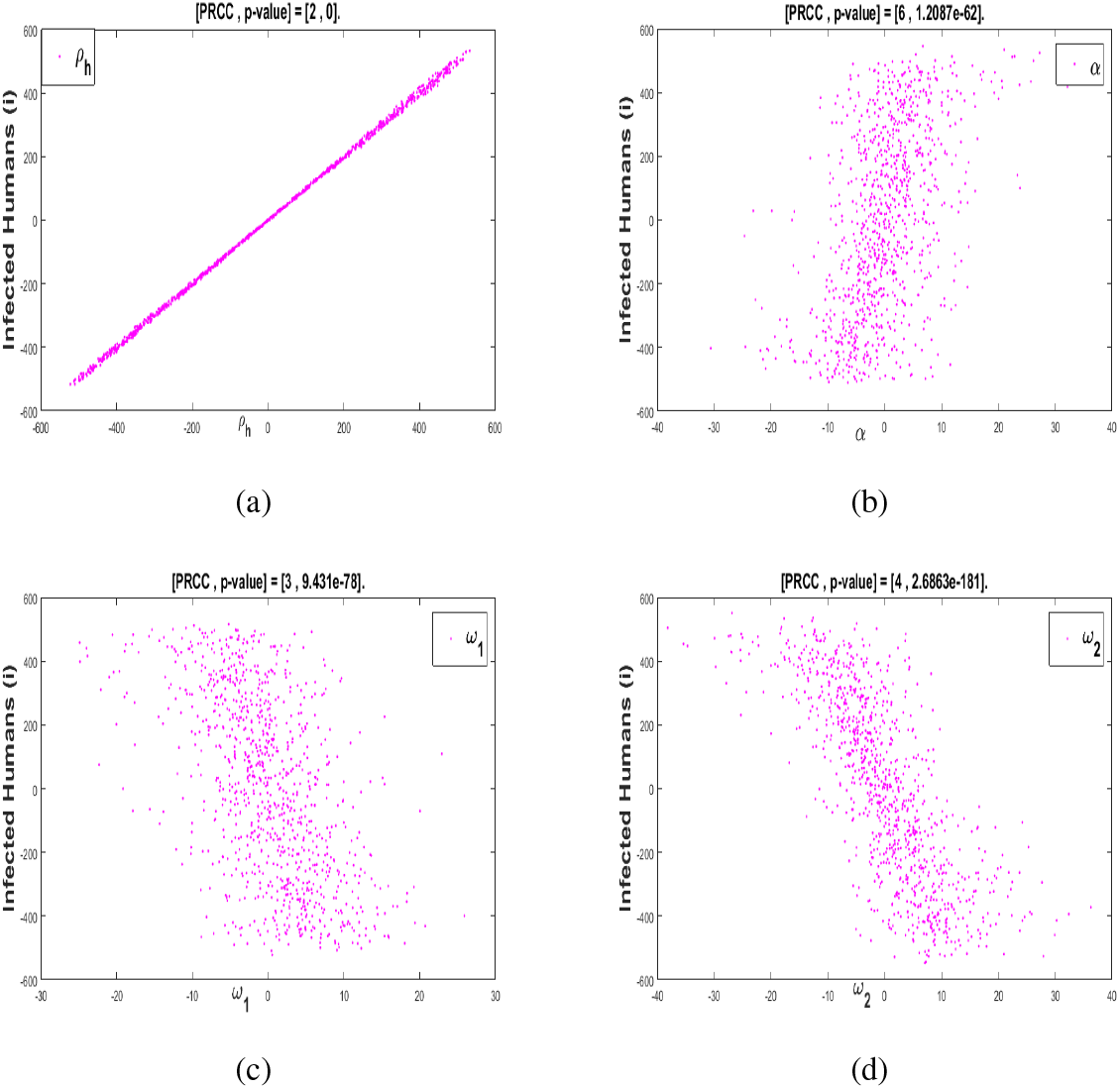
(a) and (b) Scatter plots of parameters lost of immunity rate (*ρ*_*h*_) and human recovery rate *α* with positive PRCC. (c) and (d) Scatter plots of parameters human infection rate of Listeriosis (*ω*_1_) and rate of uncontaminated food contamination by *L. Monocytogenes* (*ω*_2_) with negative PRCC.

#### 3.3.1 Varying Parameters ω_1_ and ω_2_ on the Fractions of Infected Humans

We carry out numerical simulations to support results obtained from the sensitivity analysis of the model without controls by varying parameters *ω*_1_ and *ω*_2_ on the fractions of infected humans. In Figure 4, we observe that an increase in the rate of at which humans are infected by Listeriosis (*ω*_1_) and the rate of food contamination by Listeria, (*ω*_2_) results to more fraction of infectious humans (see Figures 4(a) and 4(b)).

**Figure 4.**
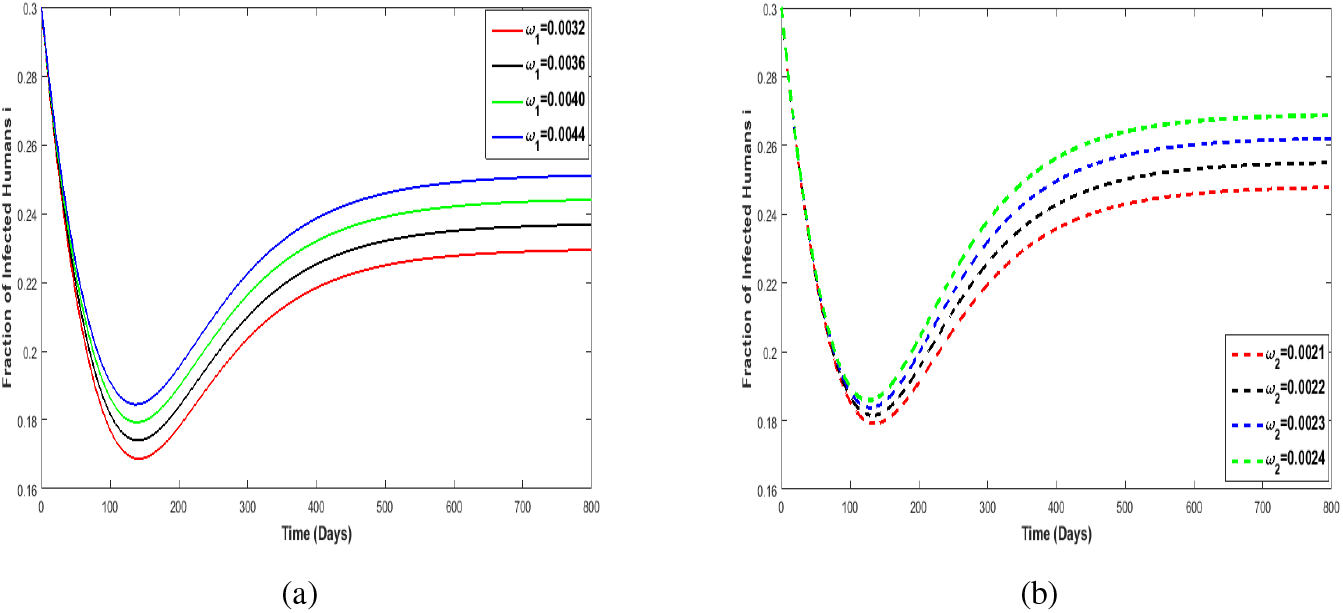
(a) Varying *ω*_1_ on *i*. (b) Varying *ω*_2_ on *i*. The value for ℜ_*f*_ is 1.5

## 4 Optimal Control Problem

In this section, we apply optimal control theory to access the preventive measures so as to reduce the spreading of the disease in a homogeneous population. Therefore, we formulate an optimal control problem with the following permissible controls variables: *u*_1_, depicting the effects of media campaigns in reducing infection; *u*_2_, controls the recovery of infected humans through the use of antibiotics; and *u*_3_ controls the removal of contaminated food products through say, the recall of food products from the retail stores or factories.

Introducing these permissible optimal control parameters *u*_1_, *u*_2_, and *u*_3_ into model equation (3), we have that:

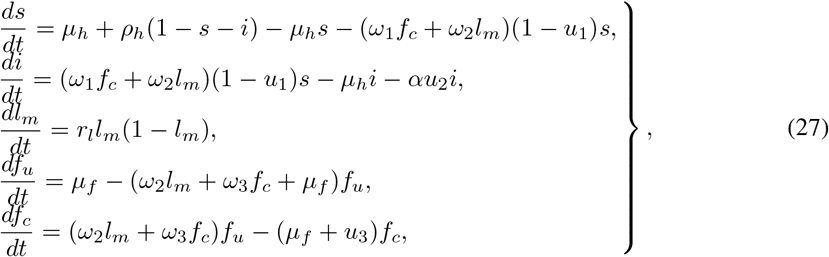

Note that all model parameters in listerirosis control models equations (27) retain the same description as defined for model system (3). The objective is to reduce the number of infected humans, hence we formulate a minimization problems with the following objective function

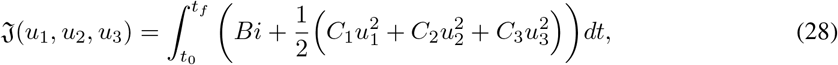

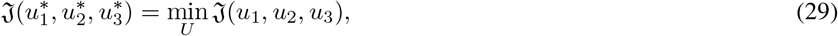

where *t*_0_ is the initial time, *t*_*f*_ is the terminal time, *B* is the weights associated with reducing the infected human class *i, C*_1_, *C*_2_, *C*_2_ are the associated cost weights with the controls *u*_1_, *u*_2_ and *u*_3_, respectively. We define the Hamiltonian function by applying the Pontryagin’s Maximum Principle Pontryagin et al. (2009) as follows

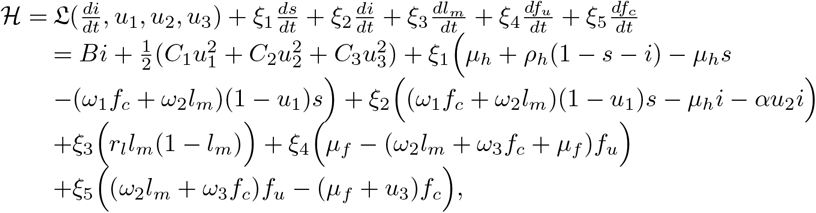

where *ξ*_*j*_, *j* = 1, …, 5 are the adjoint variables. We have the following theorem on the existence of the optimal control.

### Theorem 8

*Lenhart and Workman (2007) There exist an optimal control* 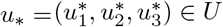 *such that*

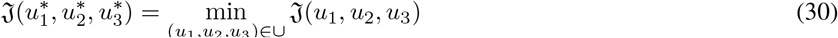

*subject to the optimal control model system (27) with initial conditions (4)*.

### 4.1 Optimality of the Model System

#### Theorem 9

*Lenhart and Workman (2007) Let s*\*, *i*\*, 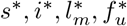, *and* 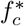 *be the solutions of the optimal control model system (27) and (28) associated with the optimal control variables* 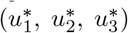. *Then there exist an adjoint system which satisfies*

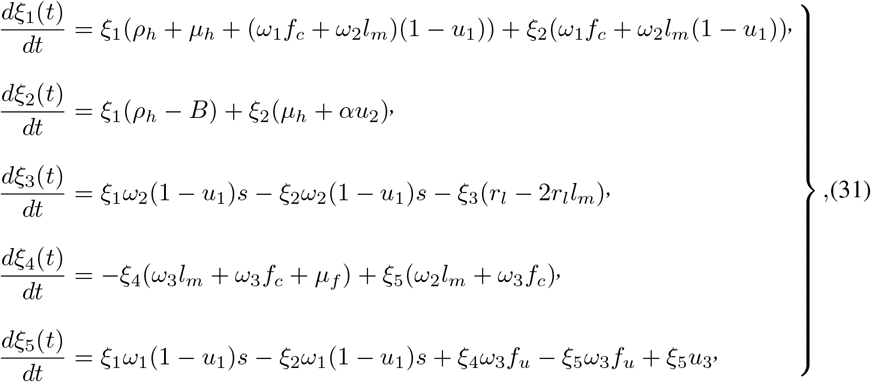

*with transversality boundary conditions denoted by*

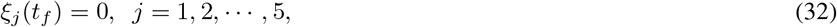

*denoted by*

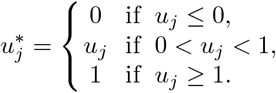

*where the permissible control functions* 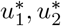, *and* 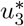, *are obtained by setting* 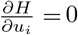, *i* = 1, 2, 3. *Thus*

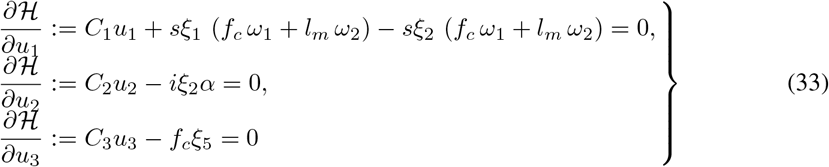

*Solving for* 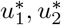, *and* 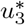 *from equation (33), result in the following permissible control solutions*

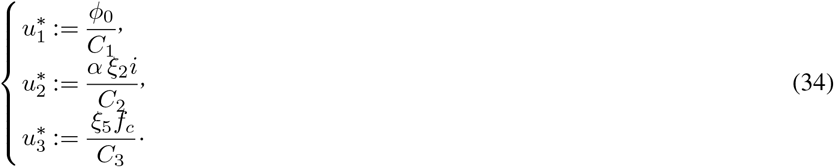

*where ϕ*_0_ = *ξ*_2_(*f*_*c*_ *ω*_1_ + *l*_*m*_ *ω*_2_)*s* − *ξ*_1_ (*f*_*c*_ *ω*_1_ + *l*_*m*_ *ω*_2_) *s Now, using the upper and lower constraints on the admissible controls* 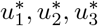, *it can be seen that the optimal controls can be characterized as:*

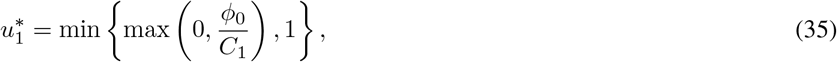

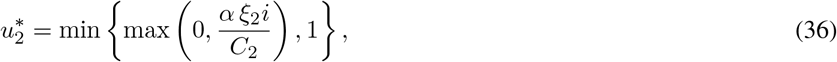

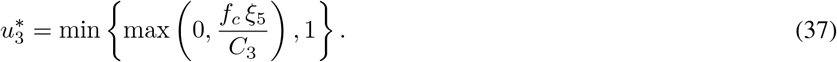

The subsequent equations (35)-(37) represent the optimality system, which has unique solution when the *t*_*f*_ is small enough and given by

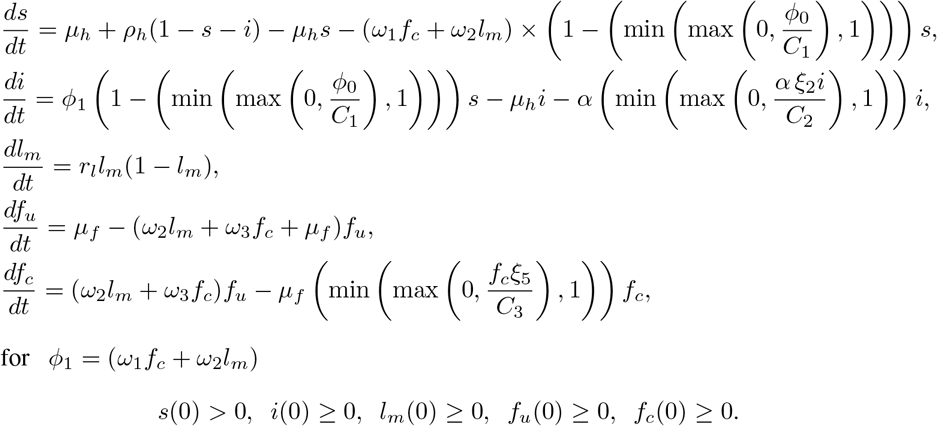

for *ϕ*_1_ = (*ω*_1_*f*_*c*_ + *ω*_2_*l*_*m*_)

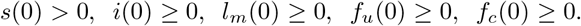

Thus, the optimality system becomes:

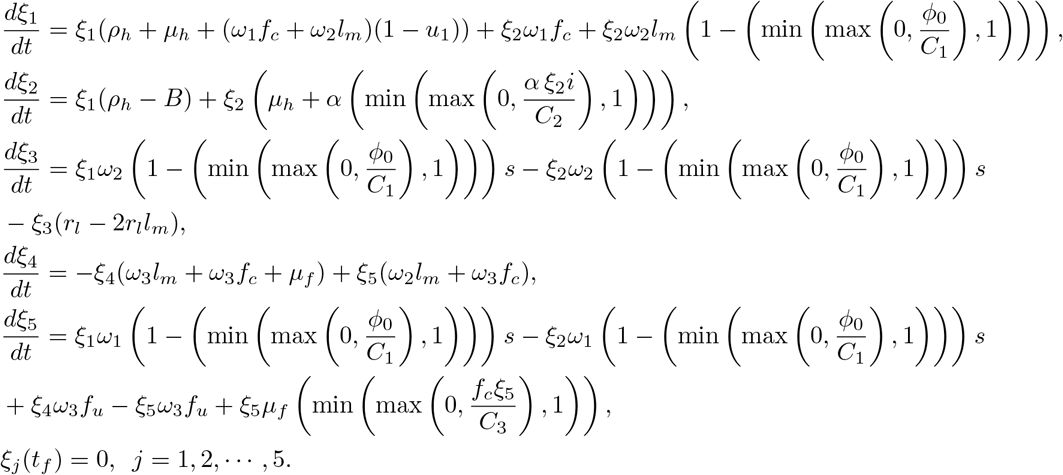

## 5 Numerical Results

The numerical solutions to the optimality system given above carried out using the Forward-Backward Sweep method Lenhart and Workman (2007) with accompanying parameter values in Tables 1 and 2. We note that, this optimal control problem is a two-point boundary-value problem with boundary conditions at time interval *t*_0_ = 0 and *t*_*f*_ = 100. The simulation is carried out for a maximum of 100 days even though the treatment of Listeriosis disease is within 30 days. In order to solve the optimal control problem, we applied Forward Runge Kutta order 4 numerical scheme to solve the model with control equations (27) and a Backward Runge Kutta order 4 numerical scheme to solve the adjoint equations given by (31). Next, we consider the permissible control values with an initial guess and find the time varying control values with an input from the computed values of the state and adjoint variables in the characterized optimal controls given by equations (34) and (35)-(37). The controls are then updated through convex combination of the control values in the previous iterations and the values from the characterized system (that means, taken the average of both the new and old control values). Further, the convergence is acquired when the values of the variables in the new iteration and the previous iteration are significantly close. The process is repeated, and iteration is stopped when the values of unknowns at the previous iteration are very close to those at the present iterations. However, for the numerical result obtained here, we have also assumed that the estimated weight of the cost media campaign, treatment and removal of food products are equal due to the type of disease (Listeriosis epidemics) in consideration, and are indicated in Table 2. In these numerical findings, we have considered combination of the following best effective possible control strategies, which are: (i) Control strategy using media campaigns, treatment and removal of food product; (ii) Control strategy using treatment only; (iii) Control strategy using treatment and removal of food product only; (iv) Control strategy using media campaigns, and removal of Food product only; and (v) Control strategy using media campaign and treatment only.

**Table 2.** Costs associated with optimal control variable used for simulations.

| Symbols | Description | Value |
| --- | --- | --- |
| $B$ | Weight associated with reducing the infected human | 1 |
| $C_1$ | Weight on the cost of media campaigns | 0.9 |
| $C_2$ | Weight on the cost of treatment | 0.9 |
| $C_3$ | Weight on the cost of removal of food products | 0.9 |

### 5.1 Control Strategy Using Media campaigns, Treatment and Removal of Food Product

In Figures 6(a) and 6(b) we observe that, in the presence of optimal control the fraction of susceptible humans increases, while the fractions of infected humans reduces over the modelling time respectively. This decline in the number of infectious individuals as seen in Figure 6(b) reflects that, the controls; media campaign and treatments has a significant impact in the human population and thus causes reduction in the number of infected humans during an outbreak. On the other hand, Figures 6(c) and 6(d) depicts that the control parameter *u*_3_ has a positive impacts on the food compartments. We notice that the in the presence of controls there is reduction in the amount of food products as time increases (Figure 6(d)) and vise versa for the uncontaminated food products (Figure 6(c)). The biological significance of this result reflects that controlling the removal of food product is essential in other to eradicate food-borne Listeriosis. Figure 5 shows the control profiles over time for the control variables *u*_1_, *u*_2_ and *u*_3_ respectively. A fascinating result can be seen in the first two control variables *u*_1_ and *u*_2_. They start at the upper bound and remains at the this upper bound, while for the control variable *u*_3_ its starts its lower bound and also remains at the lower bound within the simulation time. These results can be interpreted as follows: during an an infection period, the media campaigns needs to be implemented permanently to raise awareness in order humans to take precautionary measure to avoid been infected. Also, the removal of contaminated food products is supposed to be implemented permanently, for example, the recall of food products as implemented by South African government in the year 2018.

**Figure 5.**
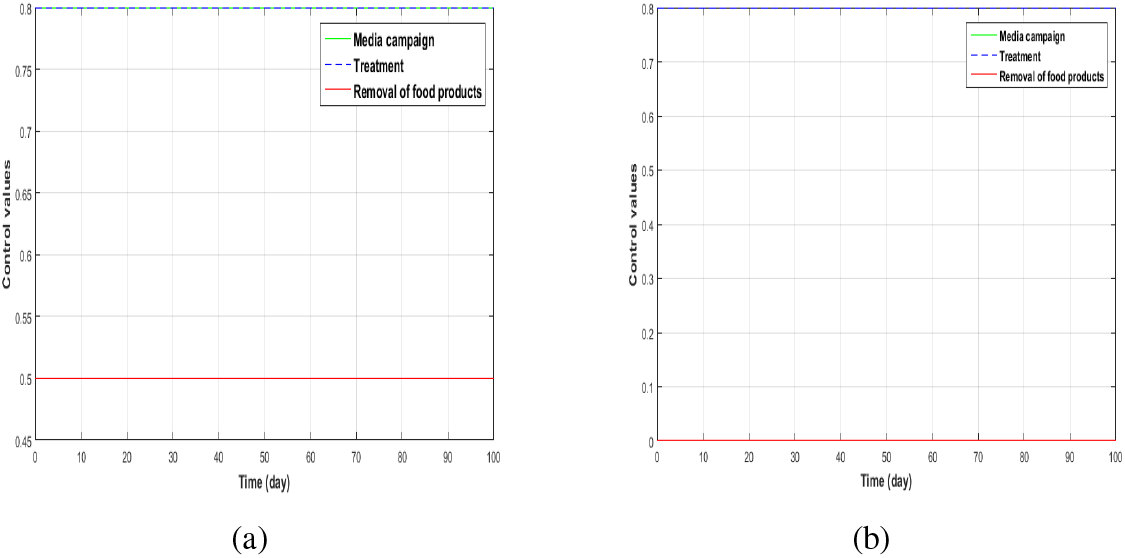
(a) Control profiles with permissible controls *u*_1_, *u*_2_, and *u*_3_. (b) Control profiles for permissible controls *u*_1_ = *u*_3_ = 0, *u*_2_ ≠ 0.

**Figure 6.**
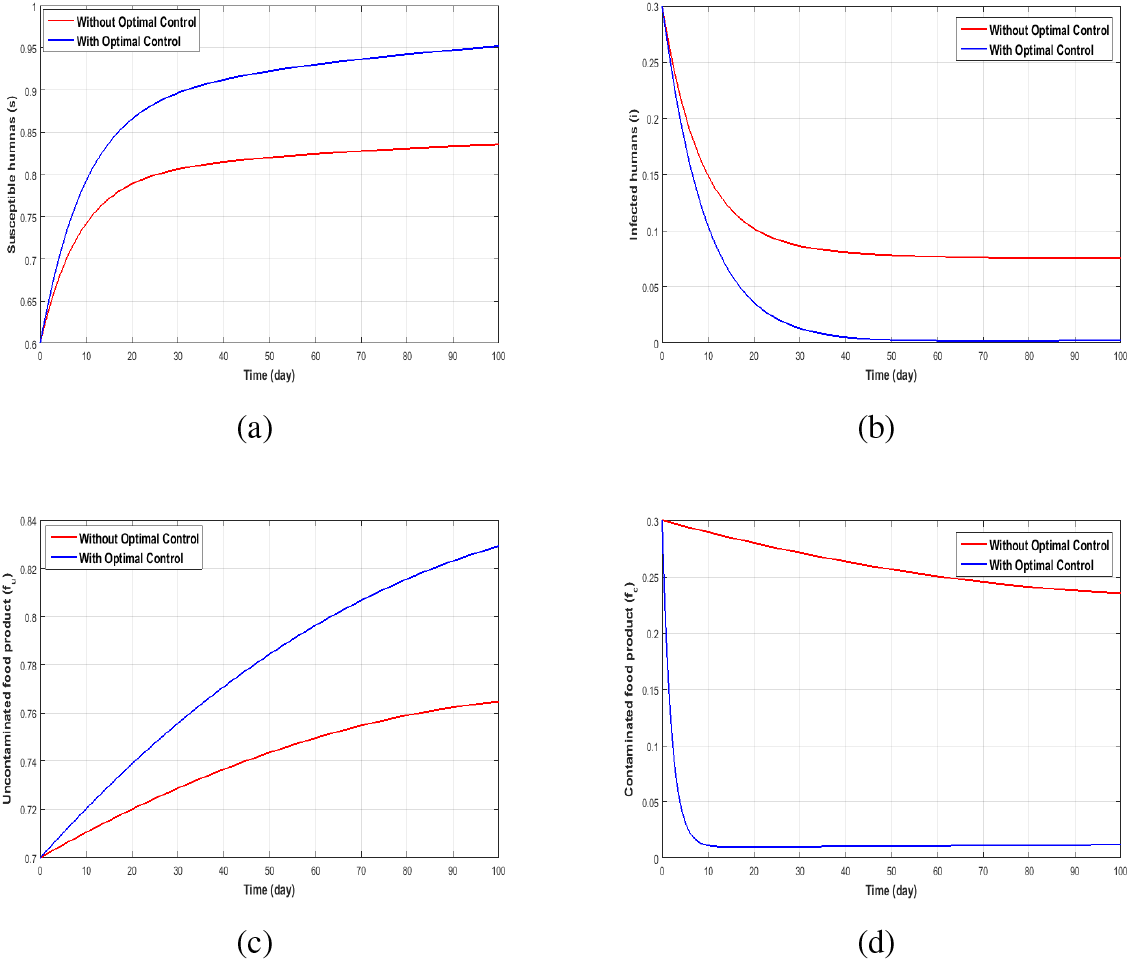
The dynamics using controls 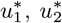 and 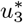 for (a) Susceptible (b) Infected (c) Uncontaminated food products (d) Contaminated food products.

### 5.2 Control Strategy Using Treatment Only

Employing treatment only as a control measure we notice that the optimal trajectories of Figure 7(a), 7(b) and Figure 7(d) do not achieve the targeted goal of minimizing the disease in the infected humans and also a reduction in the number of contaminated food, but Figure 7(c) do increase the uncontaminated food. This implies that using treatment only is not an effective way of reducing the number of infected humans in a community induced with Listeriosis.

**Figure 7.**
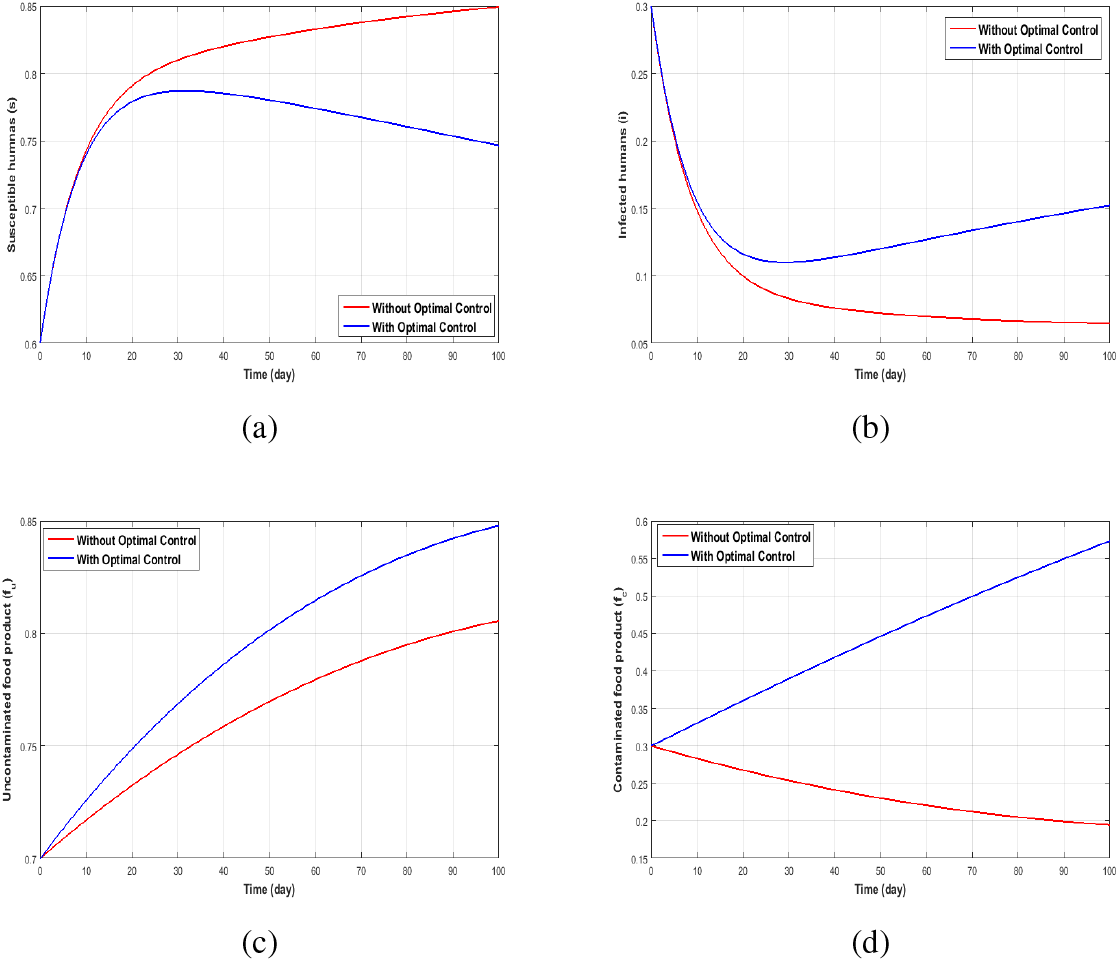
The dynamics of using permissible control *u*_2_ ≠ 0 for (a) Susceptible (b) Infected (c) uncontaminated food (d) Contaminated food.

### 5.3 Control Strategy Using Media Campaigns, and Removal of Food Products only

In Figure 9, we consider the dynamics of employing treatment and removal of food products only as a control measures. We notice that the combination of these two controls has an impact of reduction of the disease. Hence in the absence of media campaign, Listerisois can also be monitor in any community induced with the disease by the use of treatment and removal of food product only.

**Figure 8.**
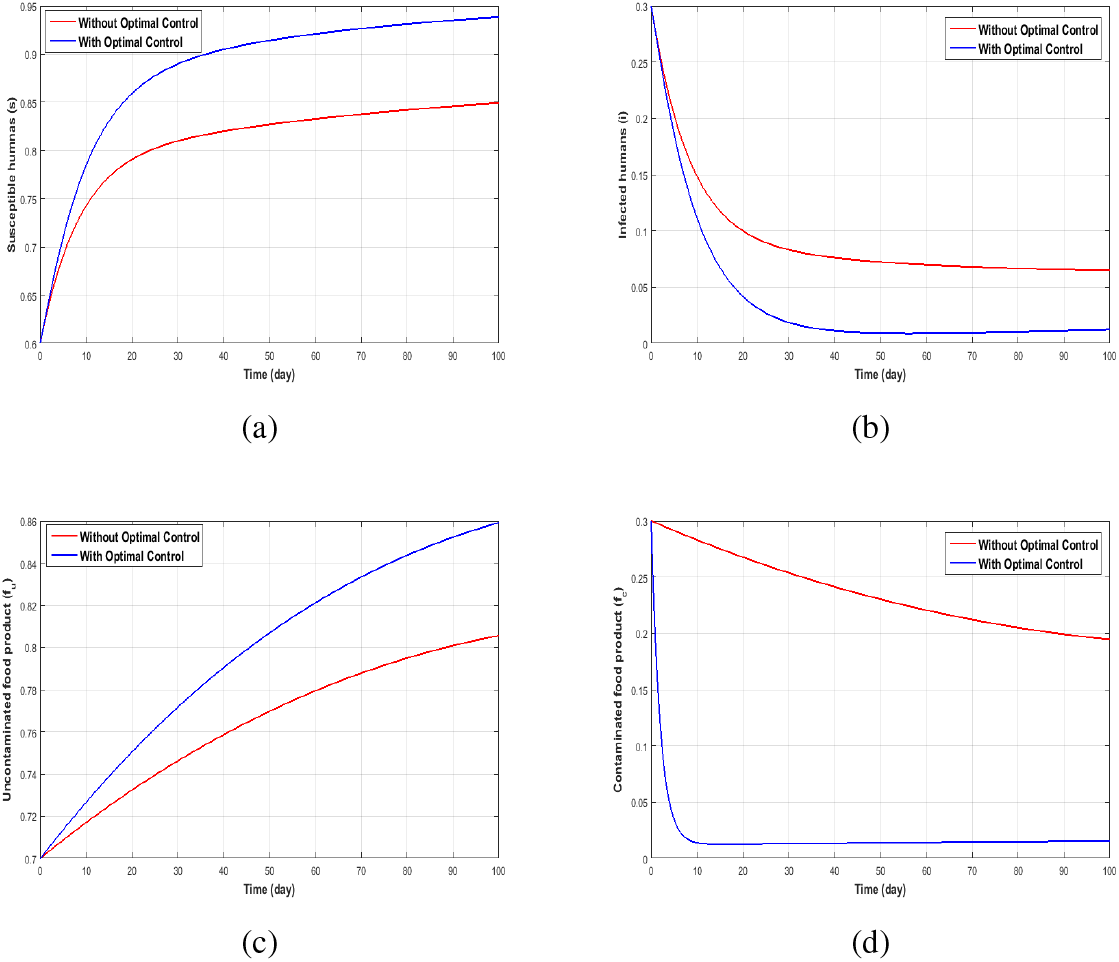
Dynamics with permissible controls *u*_3_ = *u*_2_ ≠ 0 for (a) Susceptible (b) Infected (c) Uncontaminated food (d) Contaminated food.

**Figure 9.**
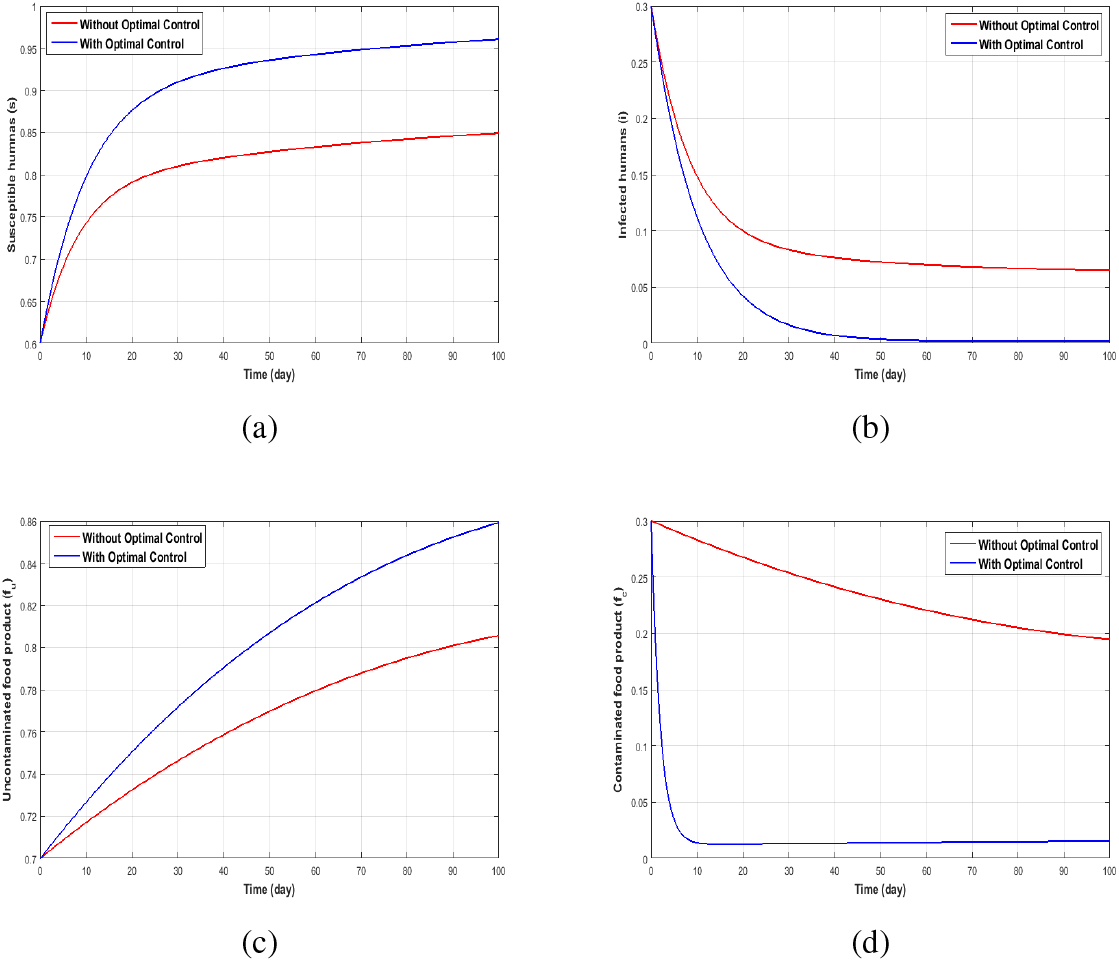
Dynamics with permissible controls *u*_1_ = *u*_3_ ≠ 0 for (a) Susceptible (b) Infected (c) Uncontaminated food (d) Contaminated food.

### 5.4 Control Strategy Using Treatment and Removal of Food Products Only

In Figure 7, we use the combination of media campaign and removal of food product to ascertain the effect of the two permissible controls. We notice that, the combination of these two controls with removal of food products play a vital role in the reduction of disease in the infected compartment faster than the possible combination of treatment and removal of food products only (subsection 5.3) and that of media campaign and treatment only (subsection 5.5).

### 5.5 Control Strategy Using Media Campaigns and Treatment Only

Finally, we study the optimal trajectories generated by using media campaigns and treatment only. We observe that the use of these control reduces the number of infections in the infected human compartment as depicted in Figure 10(b) but does not help in eradicating the disease during the entire period of the simulation period. In Figure 10(d) we can see that using these controls does not reduce the contaminated food products. This indicates that to reduce the total amount of contaminated food products it requires the combinations of all three the controls, thus media campaign, treatment and removal of food product as discussed in subsection 5.1.

**Figure 10.**
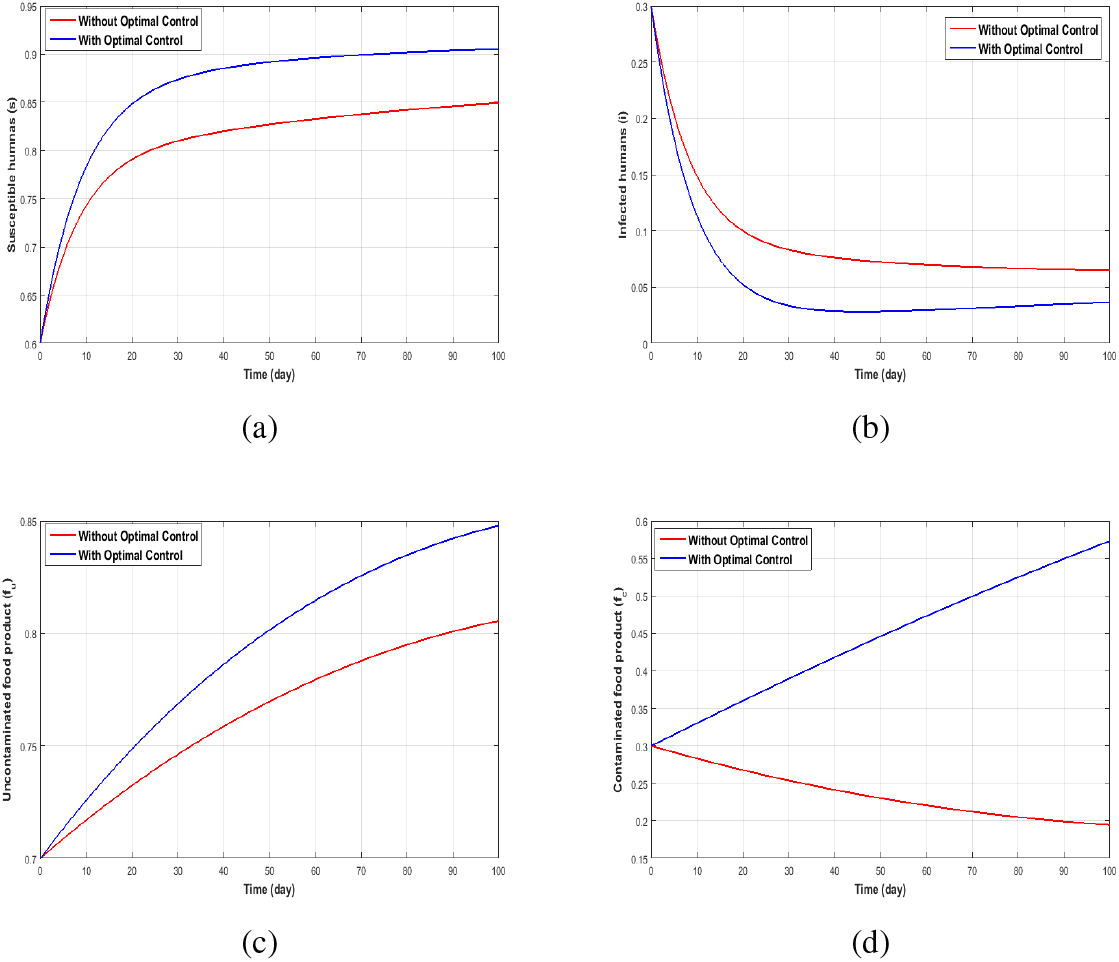
Dynamics with permissible controls *u*_2_ = *u*_3_ ≠ 0 for (a) Susceptible (b) Infected (c) Uncontaminated food (d) Contaminated food

## 6 Conclusion

In this paper, we studied the transmission dynamics of human Listeriosis disease resulting from the consumption of ready-to-eat food products, described using a set of six differential equations with the inclusion of control strategies. Mathematical analysis of the model equations without controls is presented. We found that the model has three steady states which were established to be locally asymptotically stable using the food contamination number ℜ_*f*_. The food contamination threshold is critical for the persistence or minimization of the disease. An optimal control problem was formulated with a goal to reducing the number of human Listeria infections. Numerical simulations reveal that constant implementation of media campaigns, treatment and removal of food products will be a very effective measure in the controlling and management of Listerisois in the event of an outbreak (see Figure 6). The model presented in the paper is not without shortfalls. We considered a constant human population model, implying that the long term dynamics of the model assumes the population will be constant over the modelling time, which may not be realistic. The contribution of the bacteria from the environment in an epidemic is minute. However, we argue that the bacteria in the environment can be instrumental in the growth of an epidemic. The growth function for the *L. monocytogenes* is logistic but environmental changes could impact the growth of the bacteria and hence a periodic function could better model the growth and abundance of the bacteria. Despite the shortcomings the results obtained from our findings could be used in food risk assessment to quantify the potential public health benefits on most effective control strategies to reduce food product contamination and control of human Listeria infections.

## Data Availability

None

